# Small Area Estimation of Food Insecurity in the Eastern Indo-Gangetic Plain

**DOI:** 10.1101/2021.06.03.21258287

**Authors:** Hukum Chandra, Saurav Guha, Meghana Desai, Saumyadipta Pyne

**Affiliations:** ICAR-Indian Agricultural Statistics Research Institute, New Delhi, India; Health Analytics Network, PA, USA; Public Health Dynamics Laboratory, and Department of Biostatistics, University of Pittsburgh, Pittsburgh, PA, USA

**Keywords:** Food insecurity, Calorie intake, District-level estimates, Precision, Small area

## Abstract

Achieving food security for all citizens is an important policy issue in India. While the existing data based on socio-economic surveys provide accurate estimates of food insecurity indicators at state and national level, due to small sample sizes, the surveys cannot be used directly to produce reliable estimates at the district or lower administrative levels. The availability of reliable and representative disaggregated measures of food insecurity is necessary for effective policy planning and monitoring, as food insecurity is often distributed unevenly within relatively small areas.

This article explores a small area estimation (SAE) approach to derive reliable and representative estimates of food insecurity prevalence (FIP), gap (FIG), and severity (FIS) among people in different districts of the rural areas of the Eastern Indo-Gangetic Plain (EIGP) region by linking the latest round of available data from the Household Consumer Expenditure Survey collected by the National Sample Survey Office of India as well as the latest available Indian Population Census data. District-specific food insecurity indictors such as FIP, FIG, and FIS were estimated based on a recommended threshold of per capita caloric intake of 2400 kilocalories per day, as defined by the Ministry of Health and Family Welfare, Government of India. Spatial maps showing district level inequality in the distribution of the indicators of food insecurity among the population in EIGP region are also produced. Our disaggregated estimates can provide district-specific focused insights into food insecurity to policy-analysts and decision-makers, and could thereby prove to be useful and relevant to the U.N. Sustainable Development Goal Indicator 2.1.2.

## Introduction

According to Food and Agriculture Organization of the United Nations (FAO), food security exists when all people, at all times, have physical and economic access to sufficient, safe and nutritious food that meets their dietary needs and food preferences for an active and healthy life.^1^ It is essential for the people to consume a stable diet comprising of calories and adequate nutritious food for being food secure. Conversely, food insecurity refers to a situation wherein members of society have “limited access to safe and healthy food”.^2^ In the absence of food security, nutritional deficits may lead to stunting, wasting, undernourishment, etc., that bring forth a sequalae of developmental, socioeconomic, and health consequences that impact societies in both short and long terms. In fact, it is believed that there can be no environmental security and sustainability without food security.^3,4^

Over the past 50 years, gains in agricultural productivity from the Green Revolution have translated to increased food availability. Consequently, the number of undernourished people worldwide declined from 1 billion in 1990-1992 to 795 million in 2014-2016.^5,6^ With about six-fold increase in food grain production from 50 million tonnes in 1950-1951 to about 291.95 million tonnes in 2019-2020, India has moved away from dependence on food aid to become a net food exporter country. Although food accessibility has significantly expanded, ensuring food security remains a challenge both within India and globally. As per the 2019 Global Hunger Index^7^, about 690 million people are still undernourished, 144 million children suffer from stunting and 47 million children from wasting.^7^ Indeed, the 2019 GHI ranks India in the ‘serious’ category, although it marks an improvement from the ‘alarming’ category of GHI 2000. Subsequently, the COVID-19 pandemic and its stresses to different systems have further exacerbated food insecurity for millions of people in India as well as worldwide.^8^

Research on climate change over the past decade has revealed the importance of climate pollutants (short and long-term) on monsoon patterns, precipitation changes, and increases in temperature.^9,10^ As much as 15–30% decline in production of most cereals has been projected for Africa and South Asia.^11^ In India, rice and wheat contribute to three-quarters of the country’s cereal production and their yields are susceptible to climate changes. Rainfed agriculture supports about 40% of India’s population^12^ and estimates of the past data show that changes in monsoon characteristics led to decrease in rice yields in India by 1.7% during 1960-2002.^9^ For every 1-degree Celsius increase in temperature, loss of 3.7%-14.5% in India’s wheat yields was estimated. For rice, such estimates from multiple methods predict even larger temperature impact with an average reduction of 6.6 ± 3.8% per degree Celsius.^13^

To study the complex interplay among the socioeconomic conditions, agro-ecology, and climate change, as well as their combined effects on food security of a population, few regions are as crucially important as India’s “breadbasket”, the Indo-Gangetic plain. The region comprises of a 2.5 million km^2^ fertile plain that encompasses the northern regions of the Indian subcontinent. In particular, the Eastern Indo-Gangetic Plains (EIGP) region includes the states of Uttar Pradesh (UP), Bihar, and West Bengal (WB) in India, as well as parts of Nepal and Bangladesh. In India, the EIGP region consist of 39.27 million hectares and is home to 395.19 million people, i.e., 32.64% of India’s total population (2011 census). It is among the most densely populated (700-1200 persons/km^2^) regions in the world, and has high socioeconomic vulnerability.^14^

EIGP is characterized by fertile soils with ample monsoon rainfall, continuous supply of surface and groundwater and a largely favorable climate that supports a predominantly rice-wheat cropping pattern.^15^ While UP and Bihar contribute 32% and 5.76% of the country’s total wheat production respectively, WB, Bihar, and UP contribute respectively 13.26%, 7%, and 11.75% to its total rice production.^16^ However, the food security of EIGP is potentially vulnerable to adverse effects of both anthropogenic and environmental factors. In 2012, the percentage of populations living below the national poverty line for UP, Bihar, and WB was 29.43, 33.74, and 19.98 respectively.^17^ The rates of unemployment and seasonal out-migration are relatively high among its rural populations while land holdings are generally small.

Environmental concerns of EIGP include rising temperatures, high inter-annual variability of precipitation and frequent occurrence of adverse climatic events such as droughts and increasing cyclonic activity.^18^ Annual buildups of atmospheric pollutants in intensively farmed areas may have resulted in relative yield changes of -15% or greater in this region between 2006 and 2010.^19^ The Ganga basin has severe groundwater contamination of Arsenic, which enters the food chain, especially through cultivation of rice.^20^ In fact, the disproportionately large contribution of rice production to resource use, greenhouse gases, and climate sensitivity relative to its share of monsoon cereal calorie production in India was observed.^21^ Studies have noted the importance of addressing the impacts of climate change through solutions such as diet and crop diversification, improved farming technology and issues of governance, etc.^3,21,22^

In the presence of diverse agro-ecological and biophysical conditions, availability of precise and timely disaggregate level statistics is essential for developing focused and target-oriented policies to ensure food security in EIGP. In developing countries, however, the scarcity of reliable quantitative data represents a major challenge to policy-makers and researchers.^23^ Such data, even when they exist, are often reported only at regional or state levels, and may be poorly correlated with local surveys.^24,25^ In India, Household Consumer Expenditure Survey (HCES) data collected by National Sample Survey Office (NSSO), Ministry of Statistics and Program Implementation, Government of India (GoI) is used to generate the estimates of food insecurity parameters at state and national level for both rural and urban sectors separately. However, the state and national level estimates available from this survey may not reveal the existing local disparities. In particular, estimates of food insecurity indicators are not available at the level of lower administrative units (e.g., district).

Food security presents a complex systemic challenge to policy planners, researchers, and government and public agencies. To understand it closely, one could gain key insights from statistical summaries for smaller domains of interest (or “small areas”) that are obtained by cross classifying demographic and geographic variables such as small geographic areas (e.g., districts) or small demographic groups (gender-wise, social groups, etc.) or both. However, in the existing large-scale survey data, the sample sizes of such small areas may be very small or even zero. The small area estimation (SAE) methodology provides a viable and efficient solution to this problem of small sample sizes.^26^ While SAE was recently used to obtain precise food insecurity indicator estimates for districts of Bangladesh^27^, no district-specific estimates exist for the much larger, more populous and diverse EIGP region in India.

In this study, we used SAE methodology to compute precise and representative district level estimates of the food insecurity prevalence, gap and severity among the people in rural areas of EIGP region covering the Indian states of UP, Bihar, and WB. Section 2 describes the data used for SAE as well as the study variables and model specifications. We also briefly describe the SAE methodology used in this study. The empirical results and maps are used to describe district-level inequalities in distribution of food insecurity in Section 3. Finally, we end with concluding remarks in Section 4.

## 2. Materials and Methods

### Study Area and Data Description

For this study, we used the latest round of available household consumption and expenditure survey in India, namely, the 2011-2012 HCES of the NSSO for rural areas of EIGP region which includes UP (UP), Bihar and WB and the 2011 Population Census. The de-identified survey data can be obtained from the NSSO, Government of India (http://mospi.nic.in/). Auxiliary data from the latest 2011 Population Census can be accessed freely from the Census of India website.^28^ Data obtained from these sources are then used to estimate the food insecurity indicators, viz. FIP, FIG and FIS at district level in UP, Bihar and WB. The intake of food items suggested NSSO in the 2011-12 HCES data are converted into calorie to estimate the energy intake per person per day. This survey provides information on quantity and value of more than 142 food items with a reference period of the last 30 days for a few food items and the last 7 days for the rest food items for each state separately for rural and urban areas. Average dietary energy intake per person per day in rural India is 2400 kilocalories (Kcal), as defined by the Ministry of Health and Family Welfare, Government of India.

In 2011-12 HCES, a total of 12,791 households and 67,186 persons were surveyed from the 127 districts of EIGP region. In particular, 5915 (34429), 3310 (17534) and 3566 (15223) households (persons) were surveyed from 71, 38 and 18 districts of UP, Bihar and WB respectively. The estimates of average calorie intake per capita per day for UP, Bihar, WB and EIGP region calculated from 2011-12 HCES data are 2200, 2242, 2199 and 2212 Kcal respectively. Average calorie intake for UP, Bihar, WB and EIGP region are at par with the all-India average of 2233 Kcal. The percent coefficient of variation (CV) for these estimates are 0.51, 0.74, 0.90 and 0.38 % for UP, Bihar, WB, and EIGP region respectively. The CV of state level estimates of average calorie intake are less than 1%. This evidently indicates that the higher (state/region) level estimates of average calorie intake are undoubtedly accurate. The estimates and CVs of FIP, FIG and FIS for UP, Bihar, WB and EIGP region are reported in Table 1. The estimates (and CVs) of FIP for UP, Bihar, WB and EIGP region are 0.72 (1.34%), 0.68 (2.15%), 0.73 (1.75%) and 0.71 (0.9%) respectively.

**Table 1.** The estimate and percent coefficient of variation (CV) of food insecurity prevalence (FIP), food insecurity gap (FIG) and food insecurity severity (FIS) in EIGP region.

| States | Indicators | Estimate | %CV |
| --- | --- | --- | --- |
| UP | FIP | 0.72 | 1.34 |
|  | FIG | 0.14 | 2.07 |
|  | FIS | 0.04 | 3.18 |
| Bihar | FIP | 0.68 | 2.15 |
|  | FIG | 0.12 | 3.41 |
|  | FIS | 0.03 | 5.08 |
| WB | FIP | 0.73 | 1.75 |
|  | FIG | 0.14 | 2.92 |
|  | FIS | 0.04 | 4.40 |
| EIGP region | FIP | 0.71 | 0.98 |
|  | FIG | 0.13 | 1.54 |
|  | FIS | 0.04 | 2.32 |

The FIP of Bihar (0.68) is slightly lower than both UP (0.72) and WB (0.73). Further, in UP and WB extent of FIG (0.14) and FIS (0.04) are identical whereas marginally smaller values of both FIG (0.12) and FIS (0.03) are reported in Bihar. In terms of CV, the state level estimates given in Table 1 appeared to be reasonably accurate. Table 2 summarizes district level distribution of sample size (i.e., number of persons in sample), number of food insecure person in sample (sample count) and sampling fraction in 2011-12 HCES. It is deceptive from Table 2 that the district level sample sizes and sampling fractions are quite small. Hence, it is challenging to generate the district level precise and representative estimates with associated estimates of standard errors (or CV) from this survey.^29^ This article addresses this small sample size problem in 2011-12 HCES data using SAE approach for producing the district level precise and representative estimates.

**Table 2.** District level summary of distribution of number of persons in sample (sample size), number of food insecure person in sample (sample count) and sampling fraction in 2011-12 HCES data.

| States | Summary | Sample size | Sample count | Sampling fraction |
| --- | --- | --- | --- | --- |
| UP | Minimum | 150 | 60 | 0.00017 |
|  | Average | 485 | 342 | 0.00024 |
|  | Maximum | 906 | 709 | 0.00034 |
|  | Total | 34429 | 24301 |  |
| Bihar | Minimum | 267 | 100 | 0.00014 |
|  | Average | 461 | 287 | 0.00025 |
|  | Maximum | 791 | 520 | 0.00074 |
|  | Total | 17534 | 10902 |  |
| WB | Minimum | 265 | 202 | 0.00021 |
|  | Average | 846 | 575 | 0.00026 |
|  | Maximum | 1475 | 1120 | 0.00035 |
|  | Total | 15223 | 10342 |  |

### Study Variable and Model Specifications

The target variable *Y* at the unit (person) level in the 2011-12 HCES survey data file is binary, corresponding to whether a person is “food insecure” (i.e., consumes less than 2400 Kcal per day) or not. The target is to estimate FIP (or HCR), FIG and FIS at district level for states of UP, Bihar and WB. Let *U* denotes the finite population of interest of size *N* partitioned into *D* disjoint small areas (e.g., districts here), a sample *s* of size *n* is drawn from this population with a given survey design. The set of population units in area *i*(*i* = 1,…, *D*) is denoted as *U*_*i*_ with known size *N*_*i*_ (for example, the number of people in district *i*) such that 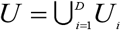 and 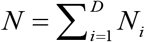. Let *s*_*i*_ denotes the collection of units making up the sample in area *i* such that 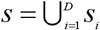 and 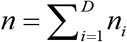. Let *y*_*ij*_ denotes the value of the variable of interest for unit *j*(*j* = 1,…, *N*_*i*_) in area *i* (for example, per capita calorie intake of person *j* in district *i*). The quantity of interest in area (or district) *i* is the food insecurity indicators *F*_*αi*_ defined as

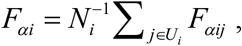

where *F*_*αij*_ = (1− *y*_*ij*_ */z*)^*α*^ *I* (*y*_*ij*_ ≤ *z*) and *z* is a preset threshold value of food insecurity (*z* = 2400 Kcal in this study), *I* (*y*_*ij*_ ≤ *z*) indicator variable for food insecure person *j* in district *i* and *α* is “sensitivity” parameter (Foster et al., 1984). The food insecurity indicators FIP, FIG and FIS are calculated by setting *α* = 0, 1 and 2 respectively.

The direct estimator (denoted by Direct) of the food insecurity indicator *F*_*αi*_ is 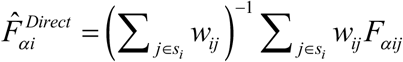, where *w*_*ij*_ denotes the survey weights associated with unit *j* in area *i*.^23^ For *α* = 0, 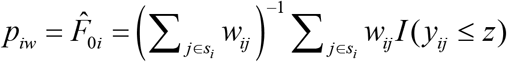 defines the survey weighted direct estimator of FIP in area *i*, 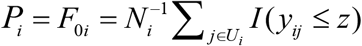. The variance of the direct estimator 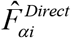 is approximated by 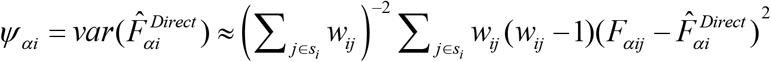. When area specific sample size is small, the sampling variance *Ψ*_*αi*_ is very large and hence the direct estimator is often unstable.

In this study, the auxiliary variables are taken from the 2011 Population Census of India. These auxiliary variables are only available as counts at district level, and so SAE methods based on area level small area models must be employed to derive the small area estimates. A range of such district level auxiliary variables is available for SAE. We choose few relevant auxiliary variables through an exploratory data analysis. We fit models with direct survey estimates of food insecurity indicators as the response variables and selected set of five auxiliary variables, viz. HH (Number of households), SC (Proportional scheduled caste population), ST (Proportional scheduled tribe population), Literacy (Literacy rate) and WP (Proportion of working population) from the 2011 Population Census as potential covariates.

In particular, a generalized linear model for FIP and the linear models for both FIG and FIS are fitted. Here, we fit a generalized linear model between district-specific sample proportions of FIP (unweighted) and set of five auxiliary variables using the glm() function in R and specifying the family as “binomial” and the district specific sample sizes as the weight. The final selected model for FIP include two auxiliary variables (ST and WP) with Akaike Information Criterion (AIC) value of -140. For this model, null deviance is 1201.9 on 126 degrees of freedom and including the two auxiliary variables (ST and WP) has decreased the deviance to 1113.2 on 124 degrees of freedom, a significant reduction in deviance. The residual deviance has reduced by 88.72 with a loss of two degrees of freedom. We also use Hosmer Lemeshow goodness of fit test to examine the fitted model (i.e. model fits depends on the difference between the model and the observed data) using the hoslem.test() function in R. The p-value of Hosmer Lemeshow goodness obtained is 0.9989. This indicates that model appears to fit well because we have no significant difference between the model and the observed data (i.e., the p-value is above 0.05).

In this fitted model, it can be noted that ST influence FIP positively, while WP has a negative effect. Further, the coefficients of ST (0.6814) and WP (−1.0305) are significant (p <0.01). This final selected model with two auxiliary variables (ST and WP) is then used for SAE to produce district wise estimates of FIP. We now fit linear models between district-specific direct estimates of FIG and FIS and set of five auxiliary variables using the lm() function in R and specifying the district specific sample sizes as the weight. Again, ST and WP are identified as two significant auxiliary variables that significantly explain the final selected models fitted for FIG as well as FIS. In the final model fitted for FIG, the regression coefficients of ST (0.19433) and WP (−0.32085) are significant (p <0.01) with R^2^ of 6%. The regression coefficients of ST (0.06176) and WP (−0.10096) are significant (p <0.01) with R^2^ of 5% for the final model selected for FIS. Hence, ST and WP are used as covariates in the SAE analysis of both FIG and FIS.

### SAE Methodology

Based on the level of auxiliary information available, the models used in SAE are categorized as area level or unit level. Area-level modelling is typically used when unit-level data are unavailable, or, as is often the case, where model covariates or auxiliary variables are only available in aggregate form. Here, we assume that the auxiliary variables are accessible at aggregate level so this article focuses on area level small area modeling. In this context, Fay– Herriot model^30^ is a widely used area level model in SAE that assumes area-specific survey estimates are available, and that these follow an area level linear mixed model with area random effects. The SAE methods based on linear mixed models for continuous data can produce inefficient and sometime invalid estimates when the variable of interest is binary.

If the variable of interest is binary in nature and the target of inference is a small area proportion (e.g., FIP), then the area level generalized linear mixed model with logit link function, also referred as the logistic linear mixed model (LLMM) is generally used. An empirical plug-in predictor (EPP) under a LLMM is commonly used for the estimation of small area proportions, see for example, Chandra et al. (2012)^31^, Rao and Molina (2015)^26^ and references therein. In this context, when only area level data are available, an area level version of a LLMM is used for SAE.^29,32^ Unlike the Fay-Herriot model, this approach implicitly assumes simple random sampling with replacement within each area and ignores the survey weights. Chandra et al. (2019)^33^ introduce an approach to model the survey weighted estimates as binomial proportions, with an “effective sample size” chosen to match the binomial variance to the sampling variance of the estimates.

In this article, we use the Fay–Herriot model^30^ to develop reliable the district level estimates of FIG and FIS. On the other hand, we consider Chandra et al. (2019)^33^ approach to model survey weighted area-specific proportions under a LLMM to produce precise the district level estimates of FIP. We briefly describe these two SAE methods. Let 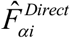 be the observed direct estimate of unobservable population parameter *F*_*αi*_ of variable of interest *y* for small area *i* (e.g., population values of FIG and FIS). In this ongoing example, districts of EIGP region are small areas of interest. Let **x**_*i*_ be the *k*-vector of known area level auxiliary variables, often obtained from various administrative and census records, related to the population parameter *F*_*αi*_. With this, the simple district (or area) specific two stage model suggested by (Fay and Herriot, 1979)^30^ is described as 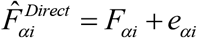 and 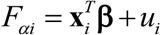. Alternatively, we can express this model as

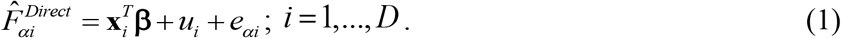

Here **β** is a *k*-vector of unknown fixed effect parameters, *u*_*i*_ ‘ *s* are independently and identically distributed normal random errors with *E*(*u*_*i*_) = 0 and 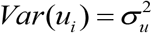, and *e*_*αi*_ ‘s are independent sampling errors normally distributed with *E*(*e*_*αi*_ | *F*_*αi*_) = 0, *Var*(*e*_*αi*_ | *F*_*αi*_) =*Ψ*_*αi*_. The two errors are of each other within and across areas (or districts). Usually, the sampling variance *Ψ*_*αi*_ is known and 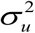 is unknown and has to be estimated. Methods of estimating 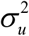 include maximum likelihood (ML) and restricted maximum likelihood (REML) under normality. Let 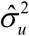 denote the resulting estimator of 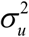 and 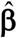 the empirical best linear unbiased estimator of **β**.

Then under (1), the empirical best linear unbiased predictor (EBLUP) estimate of *F*_*αi*_ is

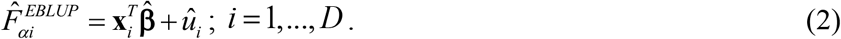

Here, 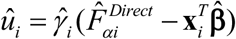, where 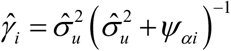 defines the shrinkage effect for area *i*. The mean squared error (MSE) estimation of EBLUP is followed from Molina and Rao (2015).^26^

The direct estimate of proportions (e.g., FIP) and area level auxiliary variables **x**_*i*_ can also be modelled by Fay-Herriot model (1) and the EBLUP estimate of small area proportions can be obtained. However, the estimate of small area proportions derived using EBLUP might be inconsistent in the sense that they might not be within the [0, 1] interval. We describe approach to model area-specific proportions under a LLMM to produce precise the district level estimates of FIP. Let 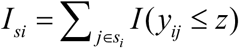 denotes the sample count (e.g., number of food insecure person in sample) in area (or district) *i*. If we ignore the sampling design, the sample count in area *i* can be assumed to follow a Binomial distribution with parameters *n*_*i*_ and *Π*_*i*_, i.e., *I*_*si*_ |*v*_*i*_ ∼ Bin(*n*_*i*_, *Π*_*i*_), where *Π*_*i*_ is success probability. Following Johnson et al. (2010)^32^ and Chandra et al. (2011)^29^, the model linking the probability *Π*_*i*_ with the covariates **x**_*i*_ is the LLMM of form

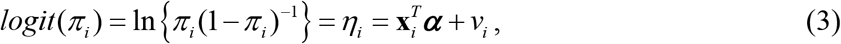

with 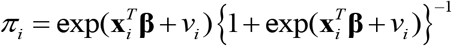. Here *v*_*i*_ is the area-specific random effect that capture the area dissimilarities with 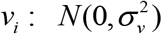 and ***α*** is the *k*-vector of regression coefficients, often known as fixed effect parameters. Under (3), a plug-in empirical predictor (EPP) of proportion (e.g., FIP) *F*_0*i*_ in area *i* is

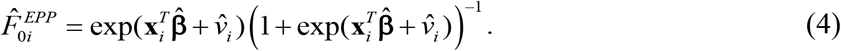

We use an iterative procedure that combines the Penalized Quasi-Likelihood estimation of ***α*** and **v** = (*v*, …, *v*)^*T*^ with REML estimation of 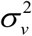 to estimate unknown parameters in (4).

The MSE estimation of EPP is adopted from Chandra et al. (2011).^29^ The model (3) is based on unweighted sample counts, and hence it assumes that sampling within districts is non-informative given the values of the auxiliary variables and the random district specific effects. Here, the survey weighted probability estimate for a district is modelled as a binomial proportion, with an “effective sample size” that equates the resulting binomial variance to the actual sampling variance of the survey weighted direct estimate for the district.^33^ In particular, in model (3) the “actual sample size” and the “actual sample count” have been replaced with the “effective sample size” and the “effective sample count” respectively to incorporate the sampling information.

A Goodness of Fit (GoF) diagnostic, which is equivalent to a Wald test, checks whether the Differences 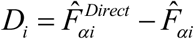 between direct and model-based estimates are statistically different, is also applied. The null hypothesis is that the direct and model-based estimates are statistically equivalent. The alternative is that the direct and model-based estimates are statistically different. This Wald test statistic is computed as 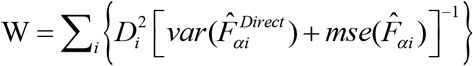. Under the assumption that 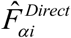 and 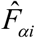 are independently distributed, which is not unreasonable for large sample sizes, the value of W can be compared with an appropriate critical value from a chi square distribution with degrees of freedom *D* equal to the number of districts.

The CV of a small area estimate 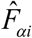 is computed as 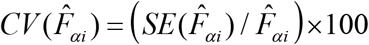, where 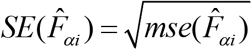 is the estimate of standard error of 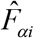 and 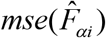 is the estimate of 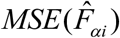. It provides a measure of relative errors and gives an indication of the precision of the model-based estimates when contrasted with the direct survey estimates. Therefore, estimates are more precise if they have smaller CVs. Conversely, the estimates with larger CVs are considered unreliable and unstable. There is, however, no single universally accepted definition of what constitutes large or small CV values.

For the 95% confidence interval diagnostic, we observe the width of the interval for the direct estimates compared to the model-based estimates.^34^ For more precise estimates, we expect the width of the confidence interval to be narrower. In addition, we consider the coverage diagnostic to assess the validity of the confidence intervals generated by the model-based SAE methods. The 95% CIs for the direct estimates should contain the “truth” approximately 95% of the time. This should also hold for the CIs surrounding the model-based estimates. We adjust both sets of intervals, so that their chance of overlapping should be 95% and count how often they actually do overlap. Assuming that the estimated coverage of the direct CIs is correct, comparing the counts to the binomial distribution provides a non-parametric significance test of the bias of model estimates relative to their precision. For accuracy of CIs for the model-based estimates, we first form adjusted 95% CIs for direct and model-based estimates using the critical values defined as 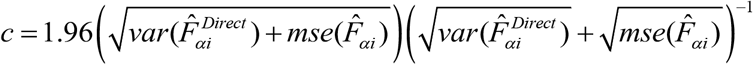. We then count the number of times the intervals do not overlap and it should be approximately 5%.^34^

Finally, we examine the aggregation property of the model-based district-level estimates of food insecurity indicators generated by SAE methods at a higher (e.g., state) level. Let 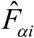 and *N*_*i*_ denote the estimate of food insecurity indicator *F*_*α i*_ and population size for district *i*. The region and state-level estimates of the food insecurity indicator *F*_*α*_ is calculated as 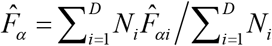, *α* = 0,1 and 2.

## 3. Results

This Section describes the district-wise estimates of food insecurity indicators (FIP, FIG and FIS) generated by our SAE methods. In particular, the EPP (4) is used to generate the district-wise estimates of FIP and the EBLUP (2) is implemented to derive the district-wise estimates of FIG and FIS. In small area applications, two types of diagnostics are executed: the model diagnostics, and the diagnostics for the small area estimates.^29,34^ The former diagnostics are examined to verify the assumptions of the underlying model, i.e., how well the working model performs when it is fitted to data. The latter diagnostics are used to provide an indication of validity and reliability of the small area estimates.

In small area models (1) and (3), the random area specific effects are assumed to have a normal distribution with mean zero and fixed variance 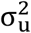. If the model assumptions are satisfied then the area or district level residuals are expected to be randomly distributed around zero. Histograms and q-q plots are also used to inspect the normality assumption. Figure 1 displays the distributions of the district level residuals (left plots), histograms (center plots) and normal q-q plots of the district level residuals (right plots) for the food insecurity indicators: FIP, FIG and FIS (top to bottom plots). The plots in Figure 1 show that the district level residuals are randomly distributed around zero. The histograms and the q-q plots also provide evidence in support of the normality assumption.

**Figure 1.**
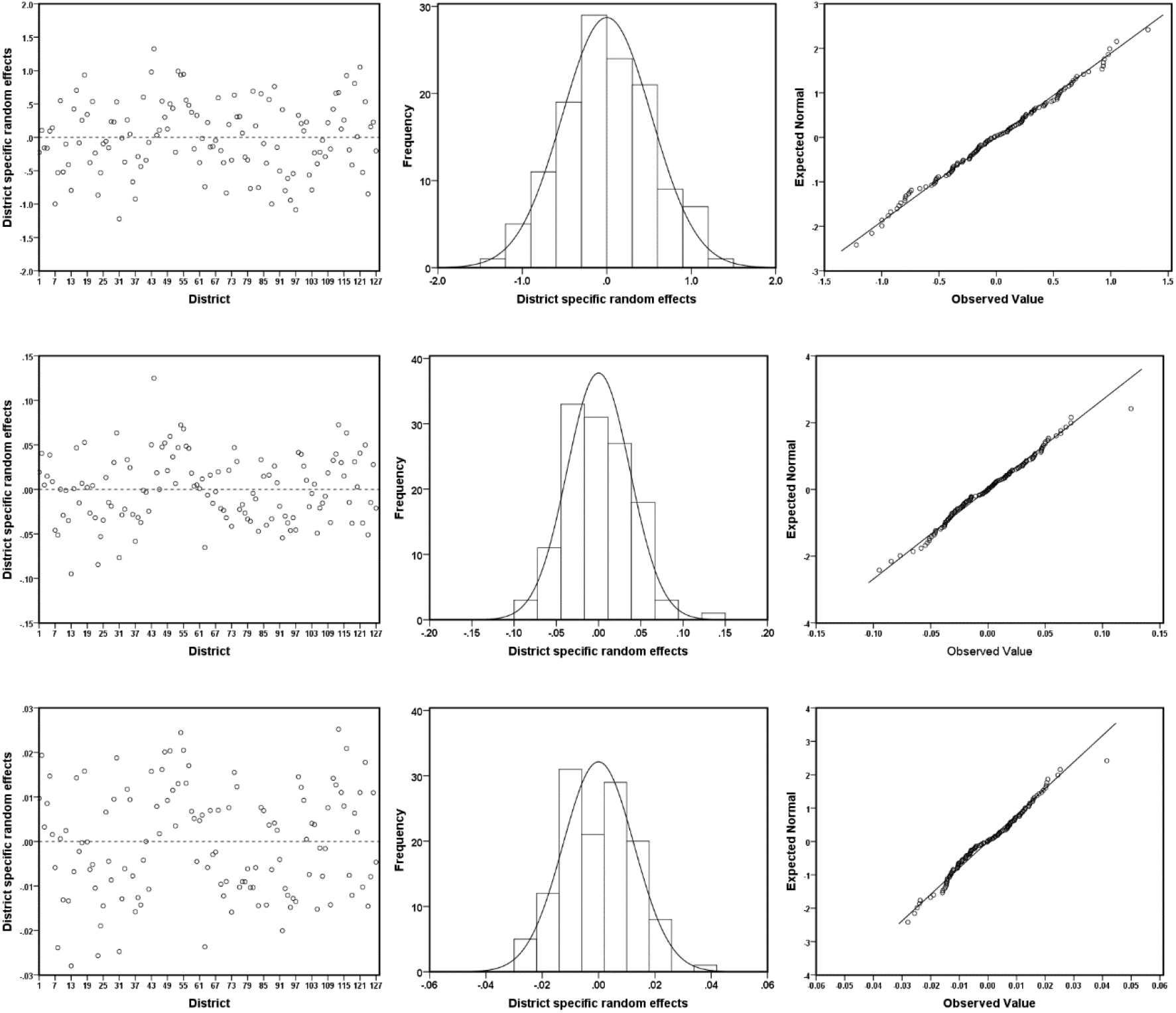
Distributions of the district level residuals (left plots), histograms of the district level residuals (centre plots) and normal q-q plots of the district level residuals (right plots) for FIP, FIG and FIS (top to bottom plots).

We also use the Shapiro-Wilk test (implemented using the *shapiro*.*test* function in R) to examine the normality of the district random effects. The Shapiro-Wilk test with p-value lower than 0.05 indicate that the data deviate from normality. The values of Shapiro-Wilk test statistics for the district level residuals, each with 127 degrees of freedom, are 0.992, 0.989 and 0.983 and p-values 0.733, 0.441 and 0.109 for models fitted with FIP, FIG and FIS respectively. In each case, the Shapiro-Wilk p-value is larger than 0.05, and hence, the district random effects are likely to be normally distributed. The model diagnostics measures evidently reveal that the normality assumptions are satisfied reasonably well with the data that we have used in this analysis.

We demonstrate three commonly used diagnostics measures for assessing the validity and the reliability of the model-based estimates of food insecurity indicators (FIP, FIG and FIS): the bias diagnostic, the percent coefficient of variation (CV) diagnostic and the 95% confidence interval diagnostic.^34^ The first diagnostics evaluates the validity and last two determine the improved precision of the model-based small area estimates. Besides this, we also explore an aggregation diagnostic where the model-based estimates are aggregated to higher level and compared with direct estimates at this level.^29^

The basic idea underpinning the bias diagnostic is that since direct estimates are unbiased, their regression on the true values should be linear and correspond to the identity line. If the model-based estimates are “close” to these true values, the regression of the direct estimates on these model-based estimates should be similar. We therefore plot direct estimates (*Y*-axis) vs. model-based estimates (*X*-axis) and we looked for divergence of the fitted least squares regression line from the line of equality.^34^ In Figure 2, we provide a bias diagnostic plots of different food insecurity indicators, defined by plotting direct estimates (*Y*-axis) against corresponding model-based estimates (*X*-axis) and testing for divergence of the fitted least squares regression line (shown by dashed line) from the line of equality, i.e., *Y* = *X* line (shown by solid line).

**Figure 2.**
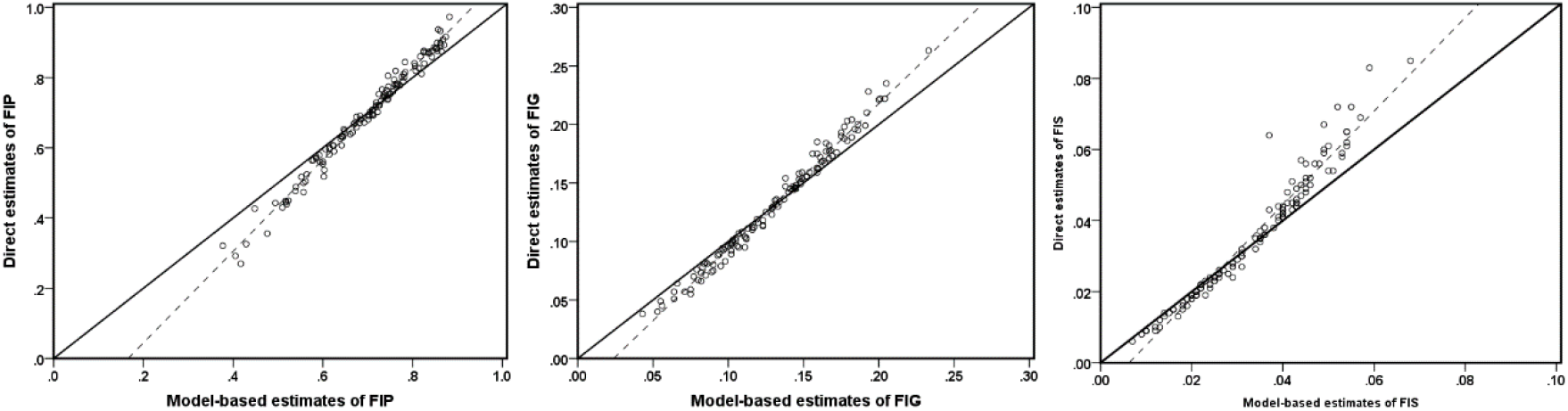
Bias diagnostic plot with *y* = *x* line (solid) and regression line (dotted) for FIP, FIG and FIS estimates: model-based small area versus direct estimates.

The bias diagnostic plots in Figure 2 clearly indicate that the model-based estimates of food insecurity indicators: FIP, FIG and FIS are less extreme when compared to the corresponding direct estimates, demonstrating the typical SAE outcome of shrinking more extreme values towards the average. The values of *R*^2^ for the fitted regression line between the direct estimates and the model-based estimates for three food insecurity indicators FIP, FIG, and FIS are 98.3, 98.5 and 95.2 per cent respectively. Similarly, the Pearson’s correlation coefficients between direct and the model-based estimates (0.992 for FIP, 0.993 for FIG and 0.976 for FIS) also support the consistency of the model-based estimates with the direct estimates.

For GoF analysis, *D* = 127, with a critical value of 154.302 at a 5% level of significance. A small value (<154.302 here) of *W i*ndicates no statistically significant difference between model-based and direct estimates. The results from GoF diagnostic are given in Table 3. The values of *W f*or the model-based estimates of FIP, FIG and FIS are 24.336, 14.788 and 18.926 respectively. These values are smaller than the 154.302, which indicates that model-based estimates are consistent with the direct estimates. In general, the bias diagnostics reflect that the model-based SAE estimates are consistent with the direct survey estimates.

**Table 3.**
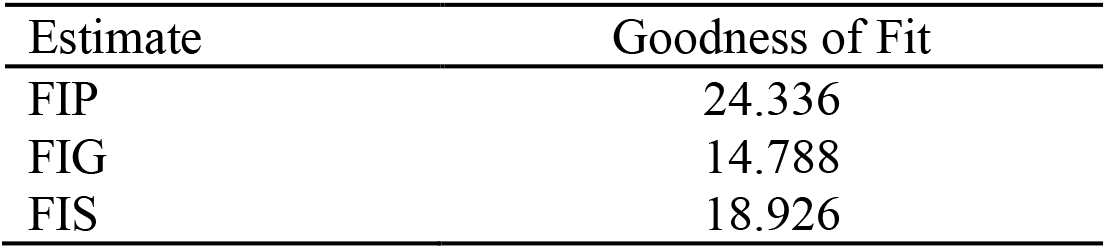
Goodness of Fit Diagnostic.

| Estimate | Goodness of Fit |
| --- | --- |
| FIP | 24.336 |
| FIG | 14.788 |
| FIS | 18.926 |

We computed the CV to compare the extent to which the model-based estimates of FIP, FIG and FIS improve in precision compared to the corresponding direct estimates. In addition, we also calculated a measure based on the ratio of the CV of the direct estimates to the model-based estimates of FIP, FIG and FIS. Figure 3 presents the district-wise values of CV (as a percentage) for model-based estimates and direct estimates of FIP, FIG and FIS in increasing order of sample sizes. Figure 4 shows boxplots of these ratios. The distribution of CV in Figure 3 indicates that in most of the districts, the CVs of the model-based estimates are significantly smaller than those of the direct survey estimates. This demonstrates that the model-based estimates are relatively more precise than the direct estimates. Further, the improvement CV is higher for the districts with smaller sample sizes as compared to the larger sample sizes. The boxplots in Figure 4 also reflect that the CV of the model-based estimates are smaller than the those of the direct estimates, implying that the model-based estimates are less variable, and hence relatively more precise than the direct estimates. Overall, the CV diagnostic measures communicate that the estimates of FIP, FIG and FIS computed from the SAE methods are more reliable and provide a better indication of the level food insecurity prevalence, food insecurity gap and food insecurity severity among the peoples in districts of EIGP region.

**Figure 3.**
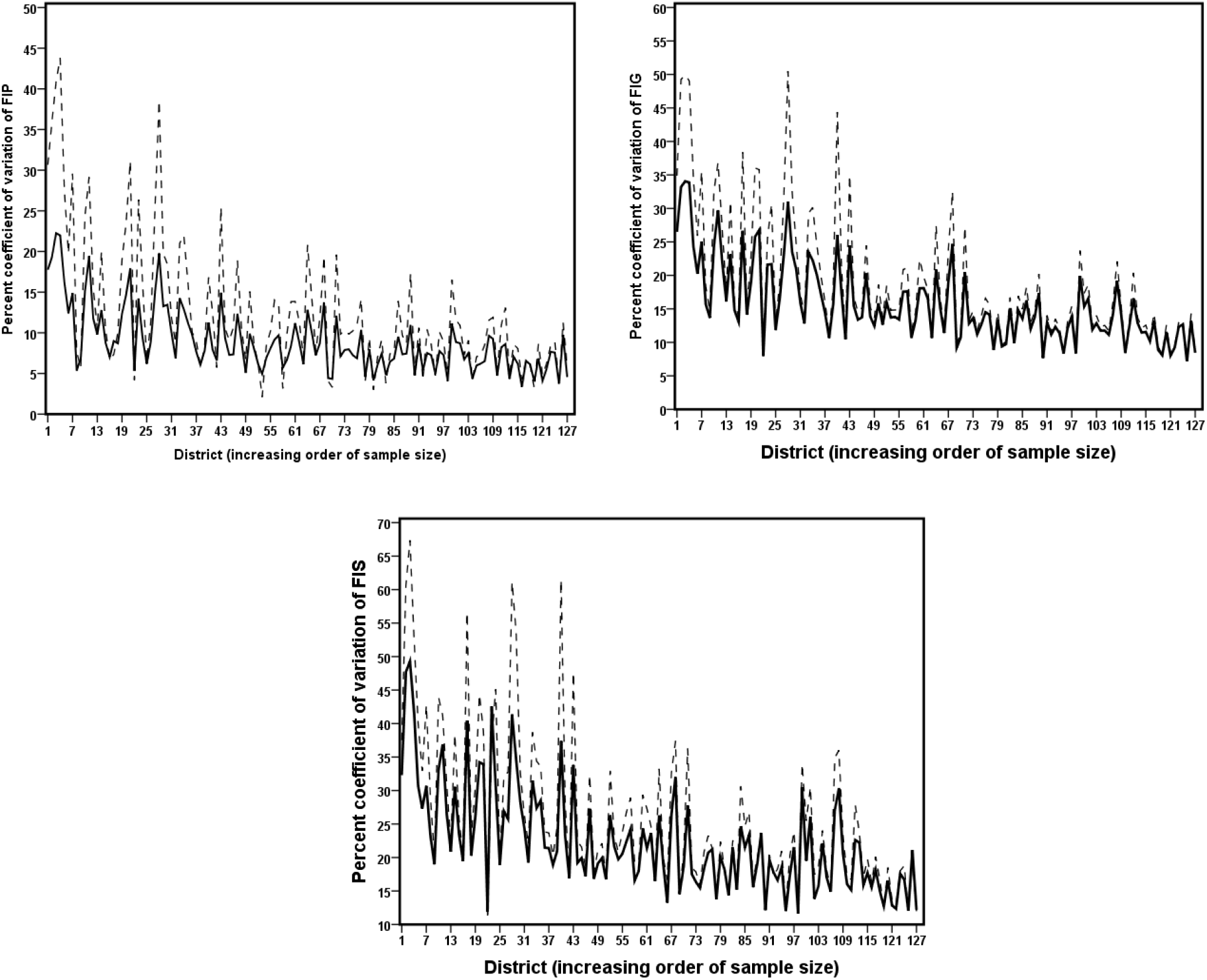
District-wise coefficient of variation (%) for the model-based small area estimates (solid line) and the direct estimates (dash line) of FIP/HCR, FIG and FIS. Districts are arranged in increasing order of sample sizes.

**Figure 4.**
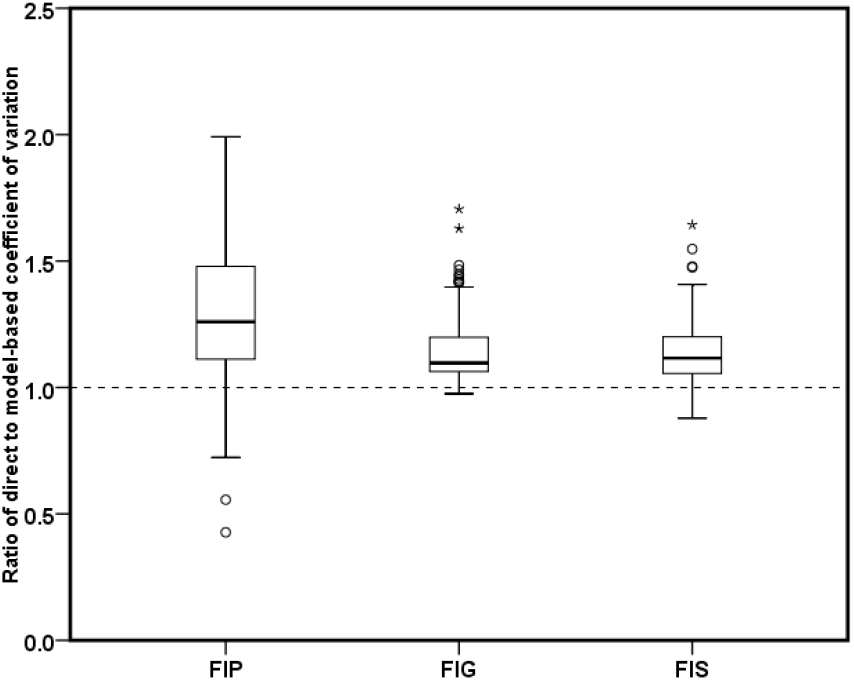
Boxplot of the distribution of the coefficient of variation of the model-based estimates compared with direct estimates of FIP/HCR, FIG and FIS.

The district-wise plots of the 95% confidence intervals (CIs) for the food insecurity indicators (FIP, FIG and FIS) generated by direct and SAE methods (EPP for FIP and EBLUP for FIG and FIS) are displayed in Figure 5. These plots show that the 95% CIs for the direct estimates are wider than the 95% CIs for the model-based estimates for the food insecurity indicators (FIP, FIG and FIS). We further note that the 95% CI for direct estimates are invalid (for example, values greater than 1.0 for FIP and negative for FIS etc.) in many districts due to large standard errors. The non-coverage percentage for FIP, FIG and FIS are 1.57%, 0% and 0% respectively, which are all under 5%. This indicates some over coverage of the model-based estimates. However, conservative CIs are not a major cause for concern.^35^

**Figure 5.**
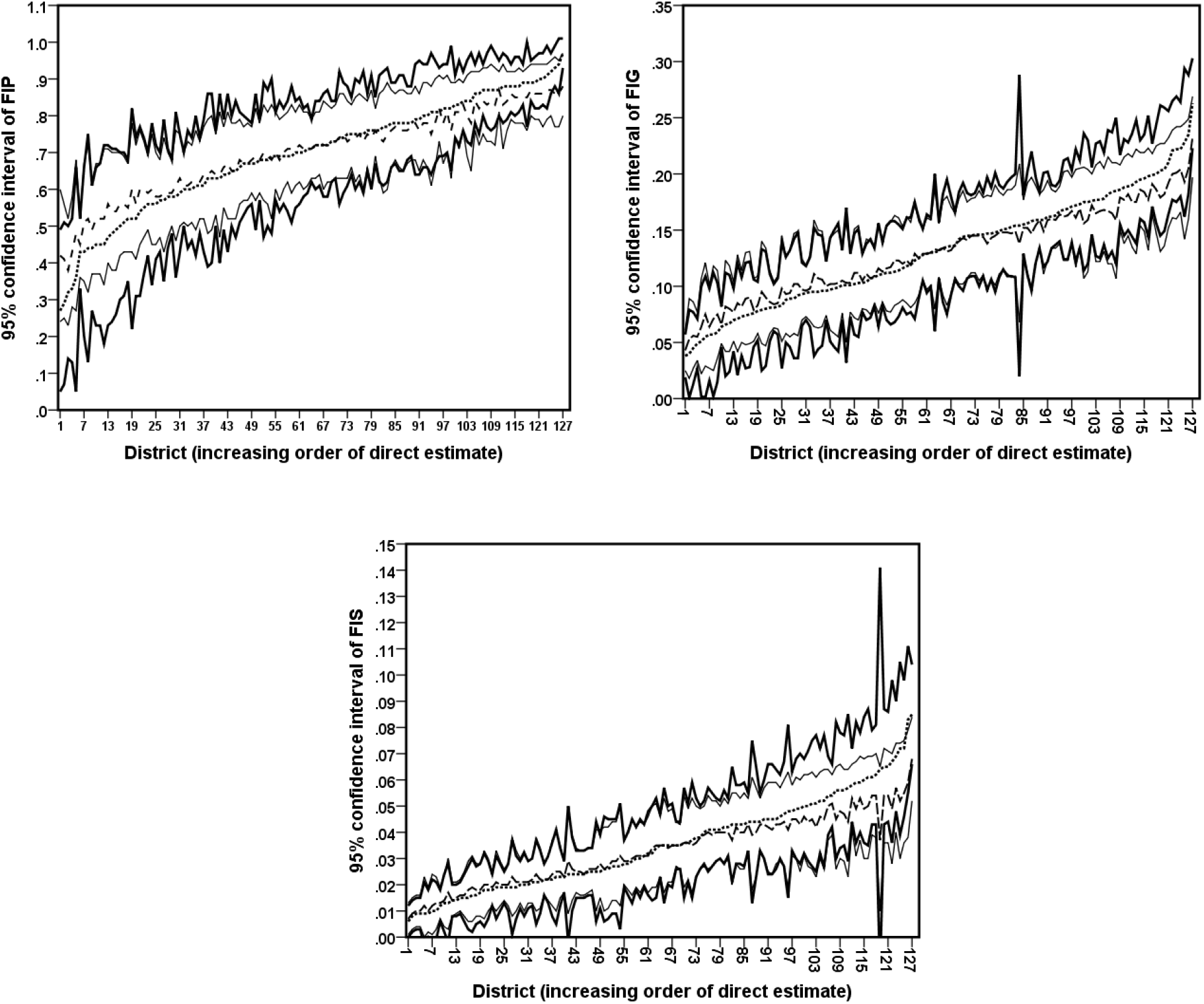
District-wise 95 percentage nominal confidence interval (95% CI) for the direct (solid line) and SAE (thin line) methods. Direct (dotted point) and model-based estimates (dash point) for the food insecurity indicators (FIP, FIG and FIS) are shown in the 95% CI.

A set of summary statistics for the direct and model-based estimates along with associated standard errors and CV of the food insecurity indicators (FIP, FIG and FIS) for 127 districts are reported in Table 4. As expected, the averages of the model-based estimates of food insecurity indicators are almost identical to those of the direct estimates but with lower variation (i.e., smaller values of standard deviation). For example, the standard deviations of FIP estimates generated by the direct and the SAE methods are 0.154 and 0.117 respectively. It is obvious that the model-based estimates of food insecurity indicators are more precise and representative than the direct estimates. To test the aggregation property, Table 5 reports the region and state level estimates of the food insecurity indicators generated by direct and SAE methods. Comparing these with the corresponding direct estimates, we note that the model-based estimates are very close to the direct survey estimates as region level as well in each of the three states.

**Table 4.**
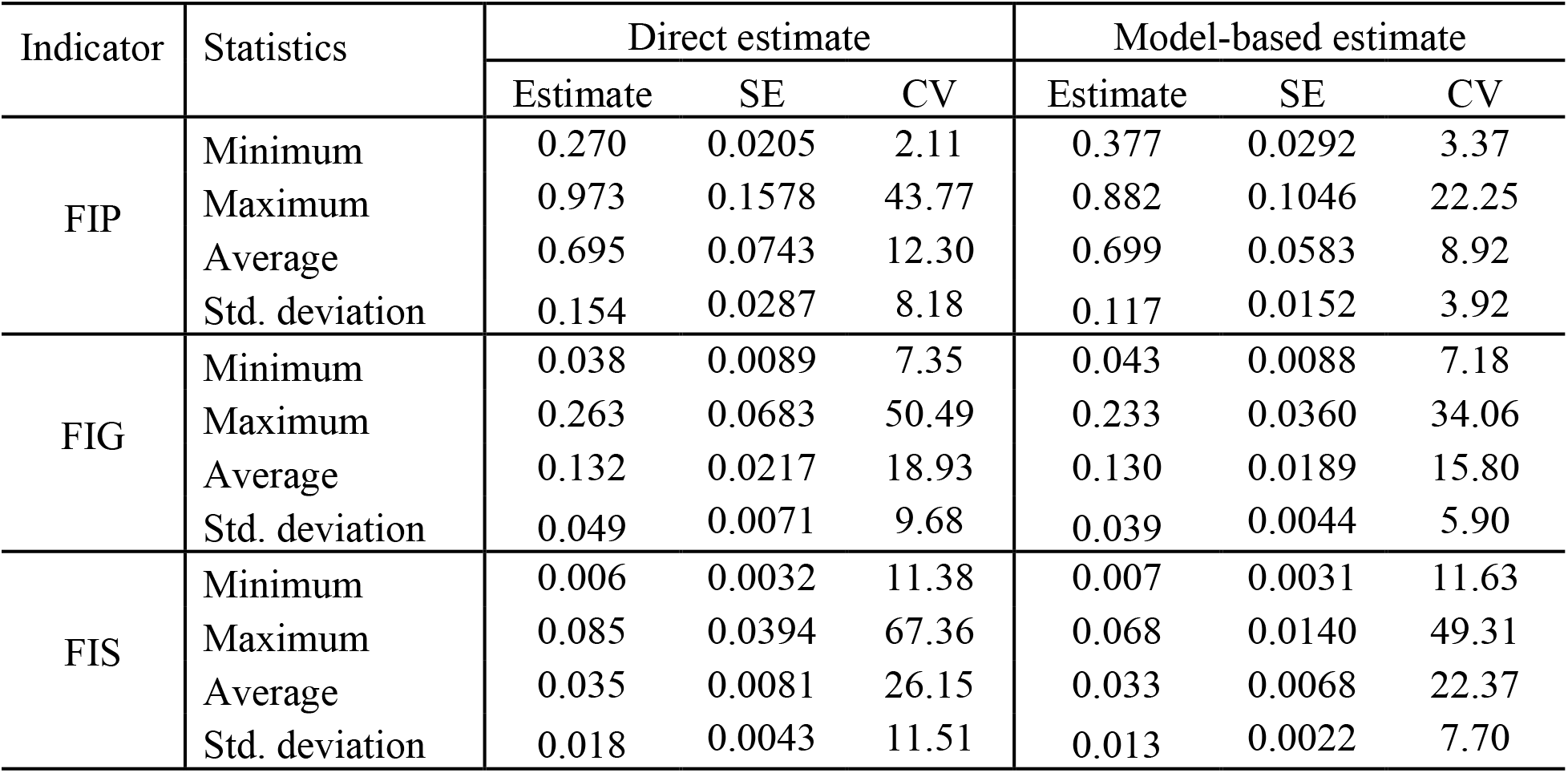
Summary distribution of direct and model-based estimates along with their standard error (SE) and percent coefficient of variation (CV) of FIP, FIG and FIS.

| Indicator | Statistics | Direct estimate |  |  | Model-based estimate |  |  |
| --- | --- | --- | --- | --- | --- | --- | --- |
|  |  | Estimate | SE | CV | Estimate | SE | CV |
| FIP | Minimum | 0.270 | 0.0205 | 2.11 | 0.377 | 0.0292 | 3.37 |
|  | Maximum | 0.973 | 0.1578 | 43.77 | 0.882 | 0.1046 | 22.25 |
|  | Average | 0.695 | 0.0743 | 12.30 | 0.699 | 0.0583 | 8.92 |
|  | Std. deviation | 0.154 | 0.0287 | 8.18 | 0.117 | 0.0152 | 3.92 |
| FIG | Minimum | 0.038 | 0.0089 | 7.35 | 0.043 | 0.0088 | 7.18 |
|  | Maximum | 0.263 | 0.0683 | 50.49 | 0.233 | 0.0360 | 34.06 |
|  | Average | 0.132 | 0.0217 | 18.93 | 0.130 | 0.0189 | 15.80 |
|  | Std. deviation | 0.049 | 0.0071 | 9.68 | 0.039 | 0.0044 | 5.90 |
| FIS | Minimum | 0.006 | 0.0032 | 11.38 | 0.007 | 0.0031 | 11.63 |
|  | Maximum | 0.085 | 0.0394 | 67.36 | 0.068 | 0.0140 | 49.31 |
|  | Average | 0.035 | 0.0081 | 26.15 | 0.033 | 0.0068 | 22.37 |
|  | Std. deviation | 0.018 | 0.0043 | 11.51 | 0.013 | 0.0022 | 7.70 |

**Table 5.** Aggregated level estimates of FIP/HCR generated by direct and model-based SAE methods in different states and EIGP region.

| Indicator | Estimator | EIGP | Uttar Pradesh | Bihar | West Bengal |
| --- | --- | --- | --- | --- | --- |
| FIP | Direct | 0.709 | 0.716 | 0.681 | 0.732 |
|  | Model-based | 0.709 | 0.715 | 0.686 | 0.726 |
| FIG | Direct | 0.134 | 0.139 | 0.122 | 0.140 |
|  | Model-based | 0.133 | 0.136 | 0.124 | 0.137 |
| FIS | Direct | 0.036 | 0.038 | 0.031 | 0.038 |
|  | Model-based | 0.034 | 0.035 | 0.031 | 0.036 |

The district-specific estimates of food insecurity indicators (FIP, FIG and FIS) along with their CVs and 95% CIs generated by the Direct and SAE methods are given in supplementary tables that are available from the authors upon request. The diagnostic measures clearly demonstrate that the model-based estimates of food insecurity indicators are more efficient, precise and representative than the direct estimates. Consequently, statistical inferences and conclusions based on the model-based estimates of food insecurity indicators are expected to offer better and effective policy decisions. In further discussion we focus and refer the model-based estimates of food insecurity indicators. Figure 6 maps the spatial distribution of FIP, FIG and FIS at district level in UP, Bihar and WB produced from the model-based estimates. Darker areas of the maps correspond to the areas of high food insecurity.

**Figure 6.**
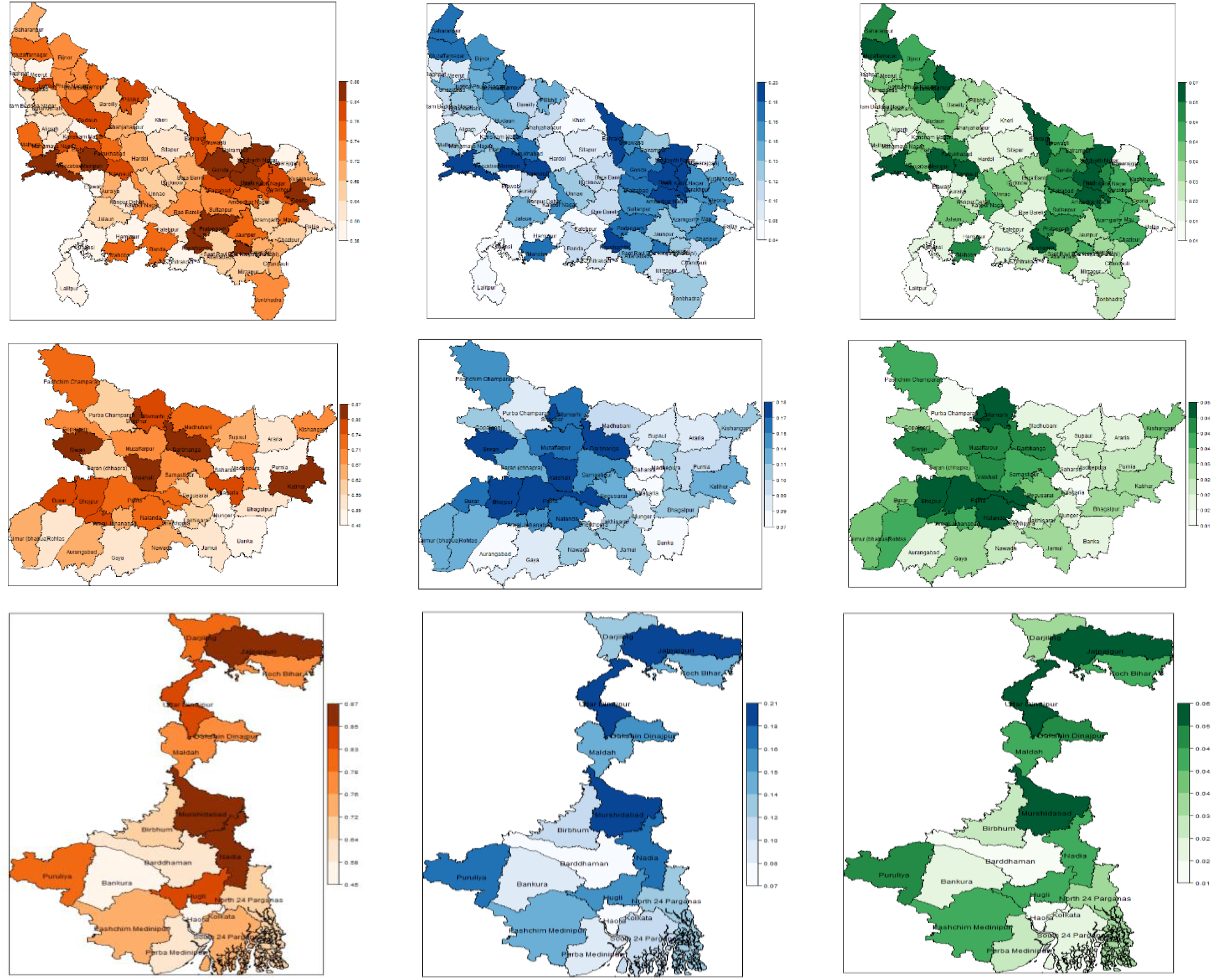
District-wise maps showing the spatial distribution of food insecurity prevalence (right), gap (center) and severity (right) generated by SAE method for the states Uttar Pradesh (Top), Bihar (Middle), and West Bengal (Bottom) in the EIGP region.

Our SAE results clearly show inequality with respect to distribution of food insecurity in different districts of EIGP region. In particular, FIP, FIG and FIS range from 37.7-88.2% (average 69.9%), 4.3-23.3% (average 13%) and 0.7-6.8% (average 3.3%) respectively between different districts in EIGP region. The disparity in distribution of food insecurity indicators between different districts within the states is also noteworthy. Food insecurity rates (FIP) between different districts in UP, Bihar and WB vary from 37.7-88.2% (average 70.3%), 40.5-87% (average 67.3%) and 44.8-86.5% (average 73.8%) respectively. Food insecurity intensity (FIG) ranges from 4.3-23.3% (average 13.3%), 7.5-17.9% (average 12.2%) and 6.6-20.5% (average 13.8%) in UP, Bihar and WB respectively. Similarly, distribution of severity (FIS) ranges from 0.7-6.8% (average 3.4%), 1.3-4.9% (average 3%) and 1.5-5.9% (average 3.6%) in different districts of UP, Bihar and WB respectively. Two emerged are these results. First, range of dissimilarities in FIP, FIG and FIS are highest in UP, as compared to Bihar and WB. Second, with larger differences in FIP, Bihar has lower level of dissimilarities in FIG and FIS than WB.

The maps in Figure 6 also demonstrate an unequal distribution of FIP, FIG and FIS in UP, Bihar and WB. In UP, the results indicate an east-west divide in the distribution of FIP, FIG and FIS. For example, in the western part of UP there are many districts with low level of FIP, FIG and FIS. In contrary, eastern part of UP have several districts with high rate of FIP, FIG and FIS. The districts with higher percentage of FIP (>80%) are Mainpuri (88.2%), Basti (87.4%), Pratapgarh (86.8%), Gonda (86.2%), Kaushambi (85.7%), Siddharthnagar (85.5%), Agra (85.4%), Sant Ravidas Nagar (84.8%), Deoria (84.3%), Pilibhit (83.5%), Sant Kabir Nagar (82.9%), Kannauj (82.5%), Ghaziabad (81.7%), Gorakhpur (81.4%), Budaun (80.7%), Bahraich (80.5%) and Faizabad (80.4%). In general, FIG and FIS status of these districts are also high except in few cases. For example, although Pilibhit, Ghaziabad and Deoria have high FIP but values of FIG and FIS index of these districts are nominal between 0.13-0.15 and 0.031-0.035 respectively. On the other hand, the districts with lower percentage of FIP (<50%) are Jhansi (37.7%), Lalitpur (41.7%), Etawah (42.9%), Mahoba (47.6%) and Meerut (49.4%).

In Bihar, except Sheikhpura (40.5%) value of FIP is greater than 50% in all other districts. But, only five districts namely Siwan (87.0%), Vaishali (86.3%), Katihar (86.2%), Darbhanga (84.3%) and Sheohar (82.5%) have percentage of FIP greater than 80%. Further, high level of FIP exists in many districts from northern part bordering with Nepal and from western part sharing border with eastern UP. For example, Pashchim Champaran (73.8%), Sitamarhi (77.9%) and Madhubani (73.4%) in northern Bihar and Siwan (87.0%), Buxar (78.1%) and Gopalganj (71.3%) districts in western Bihar. In WB, like Bihar, FIP is greater than 50% in all districts, except Howrah (44.8%) which is the neighboring district to the capital city of Kolkata. Districts with the highest FIP (>80%) are Jalpaiguri (86.5%), Murshidabad (85.5%), Nadia (85.3%), Hugli (85.2%), Uttar Dinajpur (83.7%), Puruliya (82.6%), and Darjiling (82.0%). The Jalpaiguri in the northern part, borders with Bhutan and Bagladesh in the north and south respectively, has highest FIP (86.5%), FIG (0.182) and FIS (0.053). The least vulnerable districts are Haora, Bankura and Barddhaman.

Generally, food insecurity rates, intensity and severity are mainly concentrated more in the northern and eastern parts of WB.

## 4. Discussion

The U.S. Department of Agriculture describes food insecurity as a situation of “limited or uncertain availability of nutritionally adequate and safe foods or limited or uncertain ability to acquire acceptable foods in socially acceptable ways”.^36^ This definition draws our attention to the fact that food insecurity is more than just hunger, as is recognized by the 2030 sustainable development goals (SDGs) of the United Nations, and, in particular, SDG2 that is related to ending hunger, improving food security and nutrition, and promoting sustainable agriculture.^37^ Globally, almost 2 billion people face some form of food insecurity. According to the World Food Programme, 135 million people suffer from acute hunger, and the COVID-19 pandemic is expected to double that number.

Food security and achieving the SDG2 are among the highest priorities in India. Notably, it faces a complex challenge in the form of parallel burdens of undernutrition, micronutrient deficiency as well as obesity.^38^ In 2016, 14.5% (190 million) people in India were reportedly undernourished.^39^ More recent data revealed that among Indian children of age 5 years and younger, 34.7% were stunted, 17.3% wasted, and 33.4% underweight; and among children of age 5-9 years, 21.9% were stunted, and 23% moderately-to-severely thin for their age.^40^ To summarize, one-in-three Indian children was stunted and one-in-five wasted.^40^ More than half the women of reproductive age (15-49 years) in India are anemic.^7^ Studies on long-term effects of early-life and prenatal hunger among Indian populations are ongoing.^41^

In India “food insecurity” is defined as an average calorie intake of less than 2400 Kcal per capita per day based on direct calorie intake (DCI) method.^42^ The NSSO conducts nationwide HCE surveys at regular intervals as part of its “rounds”, with the duration of each round normally being a year. The surveys are conducted through interviews of a representative sample of households selected randomly through a suitable sampling design and covering almost the entire geographical area of the country. The sampling design used in 2011-12 HCES is stratified multi-stage random sampling with districts as strata, villages as first stage units and households as second stage units. These surveys provide reliable state and national level estimates, but they cannot be used directly to produce reliable estimates at the district level due to small sample sizes. This article focuses on disaggregate level estimation and analysis of food insecurity indicators, viz., food insecurity prevalence (FIP), food insecurity gap (FIG) and food insecurity severity (FIS) using small area estimation methods.

With the policy and structural changes of the Green Revolution, countries such as India were among the first to demonstrate the gains from the high-yield varieties of cereals. While GR has substantially increased the country’s food supply over the past half century, its other more mixed outcomes include homogenization of cereal production, unsustainable resource use especially where agroecological conditions are not well-suited, and greater vulnerability to climate variations.^22^ The inter-regional differences in gains from GR reveal, especially among the EIGP districts, marked disparities in incidence of poverty, resource endowments, technology use and livelihoods.^43^ To implement its agenda of sustainable development, India currently lacks the critically essential disaggregate level measures and maps of localized food insecurity.

Towards this, the present study used the SAE approach to generate reliable and representative estimates and spatial maps of food insecurity prevalence, gap and severity among the populations in different districts of UP, Bihar and WB in EIGP region using the latest round of available data from the 2011-12 HCES and the 2011 Population Census. The results were evaluated through several diagnostics measures and revealed that the small area estimation methods offer significant gains in efficiency for generating district level estimates. Spatial maps thus produced provide key insights into the unequal distribution of incidence food insecurity, gap and severity among the different districts of these states. Our SAE approach could not only serve as a template of rigorous disaggregation for the emerging next round of the National Family Health Survey (NFHS-5) in India but also provide the much-needed precise measures for conducting systematic comparative analysis of district-wise gains or losses in food security over the past decade.

EIGP has long been studied for its relatively low productivity, poor infrastructure, limited capacity for private investment, and climate sensitivity.^44^ Our district-level estimates and spatial maps can be very effective tools in informed policy-making to address not only the challenge of food insecurity but indeed to understand the related factors that might be specific to the districts under consideration. Surely, the governmental agencies as well as various international organizations stand to benefit from our disaggregated data in formulating effective action plans to achieve the relevant SDGs. The study also underscores the potential for data-driven budget allocation and targeted welfare interventions by identification and prioritization of the districts with high food insecurity rates, intensity and severity. The COVID-19 pandemic has revealed the importance of building structural capacity with community-specific resiliency, which might have added significance in terms of regional sensitivity to climate change. For instance, formulation of policy for supporting micro, small and medium enterprises (MSMEs) in vulnerable districts could mitigate local unemployment, poverty and food insecurity. The SAE approach allows us to take a much-needed step in that strategic direction.

## Data Availability

Data was procured from freely available web resources and is mentioned in the paper.

